## Supplementary material for "Serological profiles of pan-coronavirus-specific responses in COVID-19 patients using a multiplexed electro-chemiluminescence-based testing platform": Sypplemental Figure 1

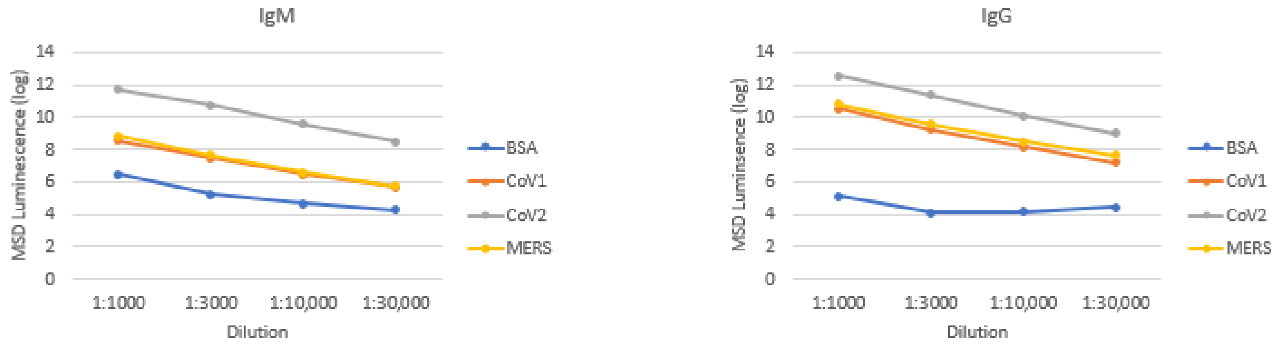


**S1 Fig: Sensitivity and specificity of assay to detect SARS-CoV-2-specific antibodies.** Serial dilutions (indicated on x-axis) of COVID19 patient samples (n=10) were testing for CoV-specific IgM (left panel) and IgG (right panel) in the 10-plex assay; plate antigens: BSA (negative control), spike protein of SARS CoV1, SARS-CoV-2, MERS CoV. Data expressed as mean luminescence signal (log).
