## Supplemental Table 1 for "Serological profiles of pan-coronavirus-specific responses in COVID-19 patients using a multiplexed electro-chemiluminescence-based testing platform"

***S1 Table:*** Age and sex of Control Subjects

| Subject ID | Age Range (y.o.) [Sex] |
|---|---|
| PG3-0061 | 60-69 [M] |
| PG3-0062 | 30-39 [M] |
| PG3-0064 | 40-49 [F] |
| PG3-0066 | 50-59 [M] |
| PG3-0067 | 30-39 [M] |
| PG3-0068 | 40-49 [M] |
| PG3-0069 | 30-39 [F] |
| PG3-0070 | 40-49 [M] |
