## Supplemental Figure 2 for "Serological profiles of pan-coronavirus-specific responses in COVID-19 patients using a multiplexed electro-chemiluminescence-based testing platform"

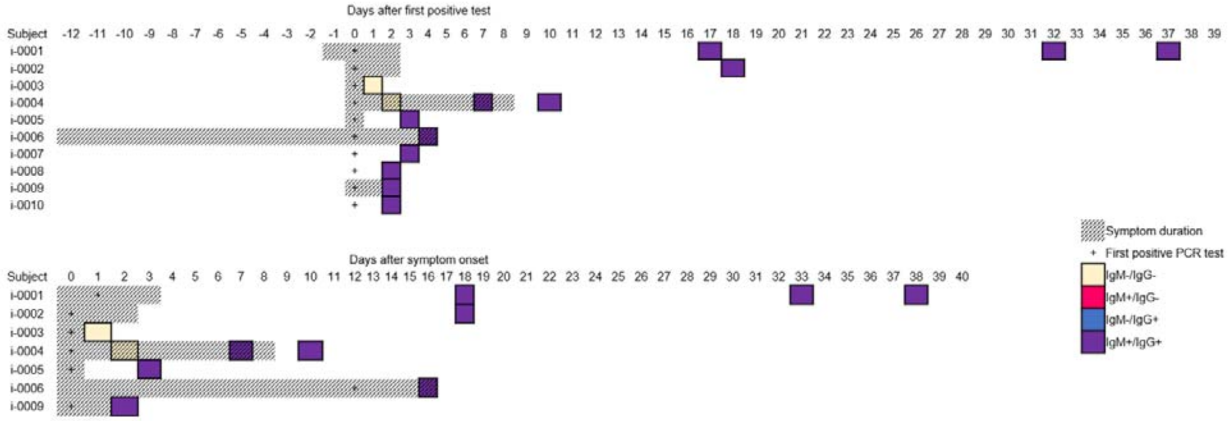


**S2 Fig: IgM and IgG seropositivity with respect to disease progression.** Seropositivity with respect to disease progression is shown for time after first postive PCR test (top) and time after sympton onset (bottom), with time of first positive test (‘+’), symptom duration (shaded), and seropositivity results (seronegative, tan; IgM+/IgG-, magenta; IgM-/IgG+, blue; IgM+/IgG+, purple).
